# A Comparative Study of Learning Outcomes between Journal Club of a Face-to-Face and an Online E-learning Course

**DOI:** 10.1101/2025.08.01.25332759

**Authors:** Hiroshi Ohara, Toshinori Ito, Keita Odanaka, Yasuhito Nobushi, Yukinaga Kishikawa, Eiji Shimanuki, Naoto Nakagawa

## Abstract

**Objective:** Evidence-based medicine requires pharmacists to critically read research papers. Journal clubs provide opportunities to develop these skills. This study compared learning outcomes between a face-to-face journal club and an online e-learning course. In evidence-based medicine, pharmacists are required to constantly read the latest clinical research papers critically and apply the knowledge gained to appropriate drug therapy. Journal clubs provide an opportunity to develop these skills. This study compared the learning outcomes of a face-to-face journal club and an online journal club using e-learning.

**Method:** A face-to-face journal club was held monthly from January to July 2019, while an e-learning course with audiovisual materials on critical appraisal was delivered from September to December 2020. Participants in both courses completed a pre- and post-course exam (5 questions) assessing critical appraisal skills and a questionnaire (five 7-level Likert items) evaluating attitudes toward critical reading.

**Results:** Both methods produced similar knowledge gains. However, the face-to-face journal club was more effective in improving attitudes toward critical appraisal, with all survey scores significantly increasing. In contrast, the e-learning group showed significant improvement in only two areas.

**Conclusions:** While both approaches enhanced knowledge, the face-to-face journal club led to greater improvement in awareness toward critical appraisal, suggesting e-learning may be less effective in this regard.

## Introduction

Reading and evaluating clinical papers as primary sources and applying them to clinical practice is important to pharmacists in the evidence-based medicine (EBM). The American Society of Clinical Pharmacists emphasizes that pharmacists should play the role of being an information source for drug treatment based on objective evidence [1]. Clinical articles provide pharmacists with tools for solving problems and questions they face in their daily work, such as handling inquiries from other healthcare professionals, planning and/or suggesting better drug therapies for each patient, and making decisions on whether to accept or reject a drug suggested by a physician. This concept is reflected in the pharmacy education in the United States, where undergraduate student pharmacists learn how to critically appraise clinical papers [2]. Journal clubs also provide pharmacists with practical training for applying findings from clinical research to their medical practice. On the other hand, our previous research has revealed that Japanese pharmacists less frequently read clinical literature than US pharmacists [3]. The Japanese pharmacists’ lack of reading habits can be attributed to the fact that methods for critical appraisal are rarely covered in their college education program. For example, knowledge of epidemiology and biostatistics, which are prerequisite for evaluating the data presented in clinical literature, is not sufficiently included in the core curriculum of the Japanese pharmacy education. Furthermore, Japanese pharmacists often have trouble in reading articles written in English. Although the need for and importance of EBM has long been recognized among Japanese pharmacists, its introduction into practice such as prescription proposals/designs remains limited.

As part of pharmaceutical informatics, there are efforts aimed at incorporating critical appraisal methods into the pharmacy student or pharmacist education curriculum [4, 5], however, it is unclear whether such efforts are widespread. A fact-finding survey on the current state of EBM education revealed that the amount of time allotted for EBM education in general pharmacist education is not sufficient in Japan [6]. In the EBM, it is important that pharmacists in the clinical environment continuously read and evaluate clinical trial articles and put the acquired information into clinical practice. Therefore, the introduction of EBM requires pharmacists’ recognition of the importance of reading articles and improving critical appraisal skills. The journal club has been a common opportunity for learning how to critically read papers and put the findings into clinical practice and the authors have been holding a journal club for students and pharmacists to spread it in Japan [4, 7]. Corona Virus Disease-19 (COVID-19) pandemic that occurred from the end of 2019 to 2023 weakened social connections and a sense of community between people. In order to keep the society going while controling the movement and physical contact between people, the utilization of information and communication technology progressed at a rapid pace. The healthcare fields were no exception, and telehealth, online medical care, online patient compliance instruction, education and meetings have become commonplace [8–11]. Based on the social situation that has forced social distancing due to the pandemic of COVID-19, we established an online e-learning program of critical appraisal skills for clinical literature using online platform as an alternative to our previously face-to-face journal club for pharmacists. The advantage of online e-learning is that it allows distance learning while maintaining social distancing, without time or geographical restrictions.

Although the purpose of the face-to-face journal club and the online e-learning course was the same: to improve critical appraisal skills, it is unclear how the different learning methods affect pharmacists’ knowledge and awareness about critical appraisal of clinical articles. In this study, we evaluated how pharmacists’ knowledge and awareness of critical appraisal changed before and after the learning courses.

## Material and methods

### Study design

The present study examined the learning outcomes between the face-to-face journal club and the e-learning course. The face-to-face journal club was held monthly from January to July, 2019. The course was for pharmacists who belonged to the Academy for Inhalation Therapy Inc. in Fukushima, Japan and who volunteered to participate.

To recruit participants for the e-learning course, we mailed a written notice on the opening of the online course and an application brochure to 788 insurance pharmacies affiliated with the Fukushima Prefectural Pharmacist Association and 113 medical institutions affiliated with the Fukushima Prefectural Hospital Pharmacist Association in June, 2020. We did not limit the number of participants and applications were accepted after participants provided their attributes (place of employment (insurance pharmacy or hospital), age, and gender). The application deadline was set on August 1, 2020 and the application could be submitted using a QR code or an URL provided in the brochure. The e-learning course was open from September to December, 2020.

### Face-to-face journal club

#### Contents and Schedule

The face-to-face course consisted of three lectures and four small group discussion sessions. The lectures, with the last author as an instructor, focused on methods for the critical appraisal of clinical literature using an instructional guide created by the authors. The article discussed in the lectures was “Extrafine inhaled triple therapy versus dual bronchodilator therapy in chronic obstructive pulmonary disease (TRIBUTE): a double-blind, parallel group, randomised controlled trial. Lancet. 2018” [12]. The articles for the small group discussion were (1) “Sher LD, et al. Fluticasone propionate and fluticasone propionate/salmeterol multidose dry powder inhalers compared with placebo for persistent asthma. Allergy Asthma. 2017” [13], (2) “Wedzicha JA, et al. Indacaterol-Glycopyrronium versus Salmeterol-Fluticasone for COPD. N Engl J Med. 2016” [14], and (3) “Randomized controlled trial of the safety and efficacy of Daptomycin versus standard-of-care therapy for management of patients with osteomyelitis associated with prosthetic devices undergoing two-stage revision arthroplasty. Antimicrob Agents Chemother. 2012” [15]. In the final session, a case of chronic obstructive pulmonary disease (COPD) was presented to the students and they were asked to discuss how to apply to the presented case the results of clinical trials on erythromycin (recommended by “Seemungal TA, et al. Long-term erythromycin therapy is associated with decreased chronic obstructive pulmonary disease exacerbations. Am J Respir Crit Care Med. 2008” [16]) and moxifloxacin (recommended by “Sethi S, et al. Pulsed moxifloxacin for the prevention of exacerbations of chronic obstructive pulmonary disease: a randomized controlled trial. Respir Res. 2010” [17]).

### E-learning Course

#### Contents and Schedule

We prepared e-learning materials that video sharing site with slide images and lecture recordings to develop reading comprehension and critical appraisal skills using the clinical trial articles. We used a free online platform to upload e-learning materials to participants and as a communication tool. The video files consisted of three parts: (1) Common English expressions in research articles, (2) Formation of English medical terms and (3) Methods of critical appraisal of clinical trial articles. The course was constructed in an appropriate order of difficulty with the easiest part first so that participants could improve their skills step by step. We selected the following articles on clinical trials: (1) “Controlled Trial of Budesonide-Formoterol as Needed for Mild Asthma. N Engl J Med. 2019,” [18], (2) “Ramucirumab plus erlotinib in patients with untreated, EGFR-mutated, advanced non-small-cell lung cancer (RELAY): a randomised, double-blind, placebo-controlled, phase 3 trial. Lancet Oncol. 2019,” [19], and (3) “Oral versus Intravenous Antibiotics for Bone and Joint Infection. N Engl J Med. 2019” [20] because they deal with diseases familiar to both insurance pharmacy pharmacists and hospital pharmacists and, therefore, knowledge bias seemed less likely to affect the learners’ comprehension. We paid royalties to the copyright holders of these articles.

#### Evaluation of learning outcomes

A knowledge-based exam to evaluate critical reading skills was set before and after the course with the following five questions: Q1. Which of the following is true about the <primary outcome= in a clinical trial?, Q2. Which of the following is true about double-blind testing?, Q3. Which of the following is correct about the double-dummy method?, Q4. Which of the following is correct regarding inclusion criteria?, Q5. Which of the following is true about the power in statistics? (See Table 1 for the details). In addition, a questionnaire concerning participants’ attitudes toward critical reading of research papers was administered to all of the participants before and after the course. It comprised the following five items: Q1. Do you agree that pharmacists need to read clinical literature?, Q2. Are you confident in your ability to read clinical trial papers?, Q3. Can you discuss the results of clinical trials with physicians and nurses?, Q4. Do you HABITUALLY read clinical trial literature?, Q5. Can you teach pharmacists how to find clinical trial literature they need? The questionnaire was scored on a 7-level Likert scale (1: strongly disagree/never, 7: strongly agree/absolutely).

**Table 1.**
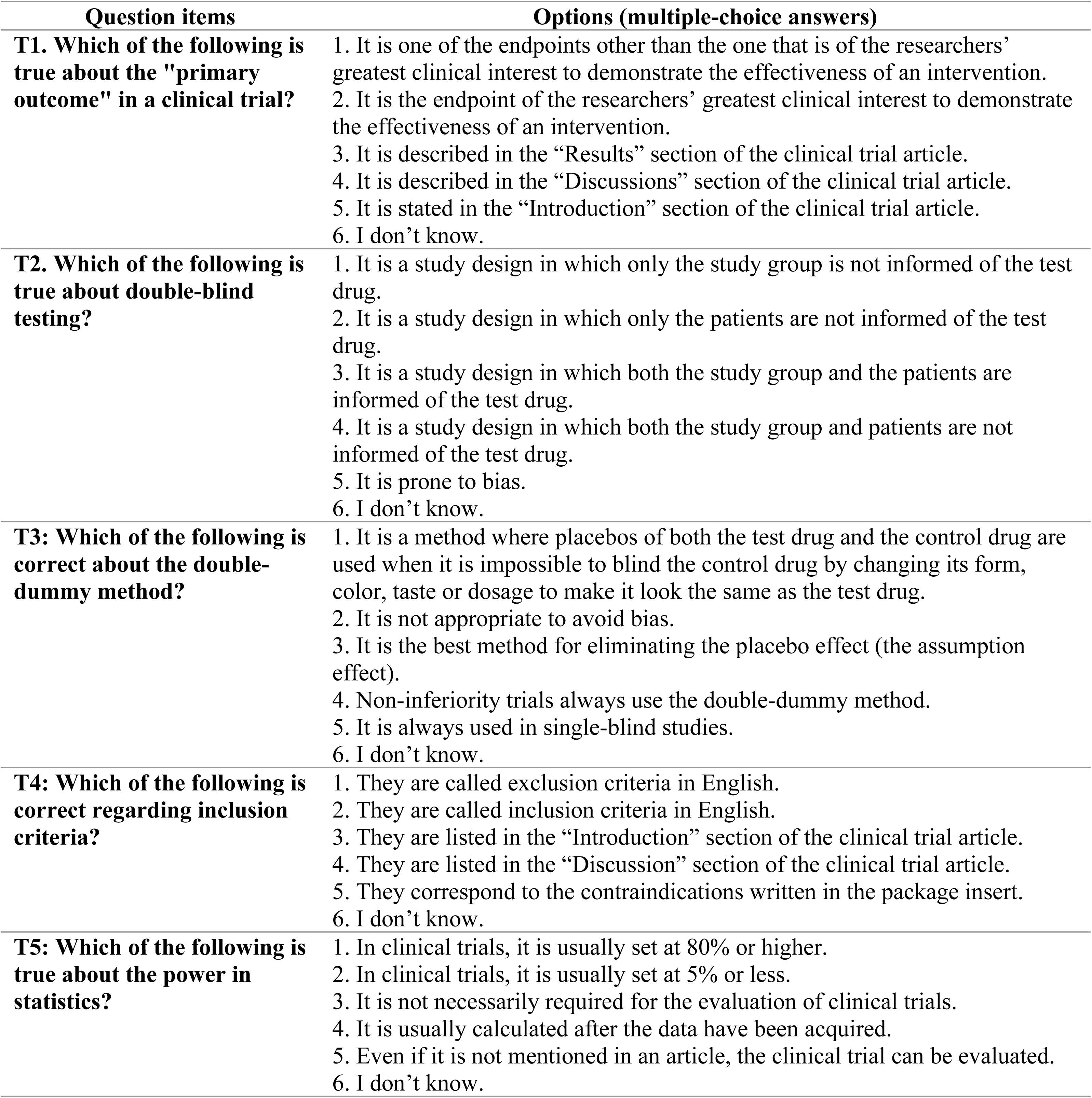
Question items and options (multiple-choice answers) about the pre-and post-tests.

| Question items | Options (multiple-choice answers) |
| --- | --- |
| **T1: Which of the following is true about the "primary outcome" in a clinical trial?** | 1. It is one of the endpoints other than the one that is of the researchers' greatest clinical interest to demonstrate the effectiveness of an intervention.<br>2. It is the endpoint of the researchers' greatest clinical interest to demonstrate the effectiveness of an intervention.<br>3. It is described in the "Results" section of the clinical trial article.<br>4. It is described in the "Discussions" section of the clinical trial article.<br>5. It is stated in the "Introduction" section of the clinical trial article.<br>6. I don't know. |
| **T2: Which of the following is true about double-blind testing?** | 1. It is a study design in which only the study group is not informed of the test drug.<br>2. It is a study design in which only the patients are not informed of the test drug.<br>3. It is a study design in which both the study group and the patients are informed of the test drug.<br>4. It is a study design in which both the study group and patients are not informed of the test drug.<br>5. It is prone to bias.<br>6. I don't know. |
| **T3: Which of the following is correct about the double-dummy method?** | 1. It is a method where placebos of both the test drug and the control drug are used when it is impossible to blind the control drug by changing its form, color, taste or dosage to make it look the same as the test drug.<br>2. It is not appropriate to avoid bias.<br>3. It is the best method for eliminating the placebo effect (the assumption effect).<br>4. Non-inferiority trials always use the double-dummy method.<br>5. It is always used in single-blind studies.<br>6. I don't know. |
| **T4: Which of the following is correct regarding inclusion criteria?** | 1. They are called exclusion criteria in English.<br>2. They are called inclusion criteria in English.<br>3. They are listed in the "Introduction" section of the clinical trial article.<br>4. They are listed in the "Discussion" section of the clinical trial article.<br>5. They correspond to the contraindications written in the package insert.<br>6. I don't know. |
| **T5: Which of the following is true about the power in statistics?** | 1. In clinical trials, it is usually set at 80% or higher.<br>2. In clinical trials, it is usually set at 5% or less.<br>3. It is not necessarily required for the evaluation of clinical trials.<br>4. It is usually calculated after the data have been acquired.<br>5. Even if it is not mentioned in an article, the clinical trial can be evaluated.<br>6. I don't know. |

#### Statistical analysis

Statistical analysis of learning outcomes between the face-to-face course and the online course was performed using the Wilcoxon rank sum test with a continuity correction. In addition, the Student’s T-test was used to conduct statistical analysis of exam scores between the two. The difference was considered statistically significant when the risk rate was less than 5%. Additionally, the effect size (Cohen’s d) was calculated using EZR (“Easy R”) (Jichi Medical University Saitama Medical Center, Saitama, Japan), a front-end application for the programming language R (The R Foundation for Statistical Computing) [21], and G*Power software [22]. In Brief, the power was calculated with the EZR, then the effect size was calculated with G*Power. The effect size of 0.20, 0.50, and 0.80 was interpreted as indicative of small, medium, and large effect, respectively [23].

#### Ethical Approval

The study was conducted with the approval of the Ohu University ethics review board in compliance with the “Ethical Guidelines for Medical Research for Humans” (approval number: 294). The participants’ responses to pre- and post-course exams and questionnaire surveys were considered their consent to participate in the study.

## Results

The background information on participants is shown in Table 2. There were no significant differences in gender or length of career between the two groups.

**Table 2.** Background information of participation.

| Method | Face-to-face | e-learning | P value* |
| --- | --- | --- | --- |
| **Number of Participants** | 18 | 25 |  |
| **Gender (Female/Male)** | 6 / 12 | 10/ 15 | 0.755 |
| **Career as pharmacist**<br>**(years)** |  |  | 0.896 |
| **1 - 5** | 2 | 6 |  |
| **6 - 10** | 4 | 4 |  |
| **11 - 15** | 4 | 6 |  |
| **16 - 20** | 3 | 3 |  |
| **21 -** | 3 | 5 |  |

379 \*: Fisher exact test

Eighteen pharmacists participated in the face-to-face journal club but two of them did not take the exams. On the other hand, 25 pharmacists were enrolled in the online e-learning course and one of them did not respond to the survey. The number of participants who responded to the post-course exam and survey was 12 and 13, respectively, where all the 12 dropouts of the post-course survey were included in the 13 dropouts of the post-course exam. The dropout rate was 52% (13 divided by 25).

Results of the exams regarding critical appraisal are shown in Figure 1. The median score of the post-course exam significantly exceeded that of its pre-course counterpart both in the face-to-face journal club (*P* = 0.043, *d* = 0.706) and in the e-learning course (*P* = 0.006, *d* = 0.703) as shown in Figure 1(a) and (b). The score of the pre-course exam (Figure 1(c)) was significantly higher in the face-to-face journal club than in the e-learning course (*P* = 0.001, *d* = 1.268) whereas the score of the post-course exam (Figure 1(d)) was not significantly different between the two groups, but the effect size was medium (*P* = 0.104, *d* = 0.660).

**Figure 1.**
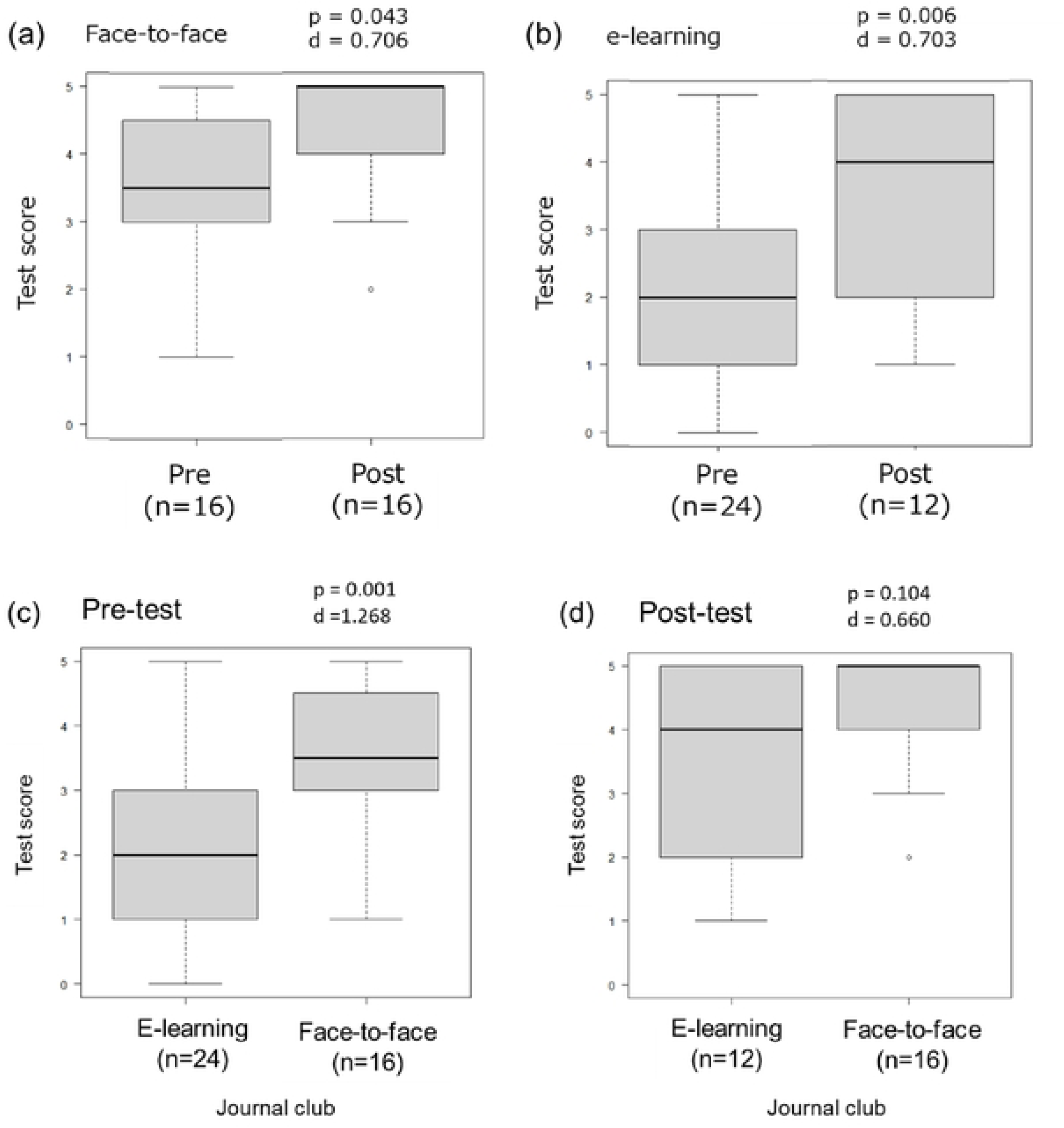
Results of exams on critical reading skills. (a) Comparison of scores between pre-course and post-course exams in the face-to-face group (b) Comparison of scores between pre-course and post-course exams in the e-learning group (c) Comparison of pre-course exam scores between the face-to-face and the e-learning groups (d) Comparison of post-course exam scores between the face-to-face and the e-learning groups

Results of the questionnaire surveys are shown in Figure 2. The score of “Q1. Do you agree that pharmacists need to read clinical literature?” was relatively high in the pre-course survey in both groups. However, the score of the post-course survey significantly increased only in the face-to-face group (face-to-face: *P* = 0.049, *d* = 0.681, e-learning: *P* = 0.478, *d* = 0.095) (Figure 2(a-b)). The score of “Q2. Are you confident in your ability to read clinical trial papers?” was significantly higher in the post-course survey in both groups. More specifically, the median score was 1 in the pre-course survey and 4 in the post-course survey in the face-to-face group (Figure 2(c), *P* < 0.001, *d* = 1.583), while it was 1 and 3 respectively in the e-learning group (Figure 2(d), *P* = 0.001, *d* = 1.184). Regarding “Q3. Can you discuss the results of clinical trials with physicians and nurses?”, we found a significant increase in the score in the face-to-face group (Figure 2(e), *P* < 0.001, *d* = 1.184), but not in the e-learning group (Figure 2(f), *P* = 0.196, *d* = 0.264), as in Q1. Similarly, for “Q4. Do you HABITUALLY read clinical trial literature?”, there was a significant increase in the score in the face-to-face group (Figure 2(g), *P* < 0.001, *d* = 0.884), but not in the e-learning group (Figure 2(h), *P* = 0.224, *d* = 0.388). As for “Q5. Can you teach pharmacists how to find clinical trial literature they need?”, we found a significant increase in the score in both groups (face-to-face: *P* < 0.001, *d* = 1.227, e-learning: *P* = 0.028, *d* = 0.645) (Figure 2(i-j)). To sum up, all scores significantly increased in the face-to-face group, while significant increase was only observed in two questions in the e-learning group.

**Figure 2.**
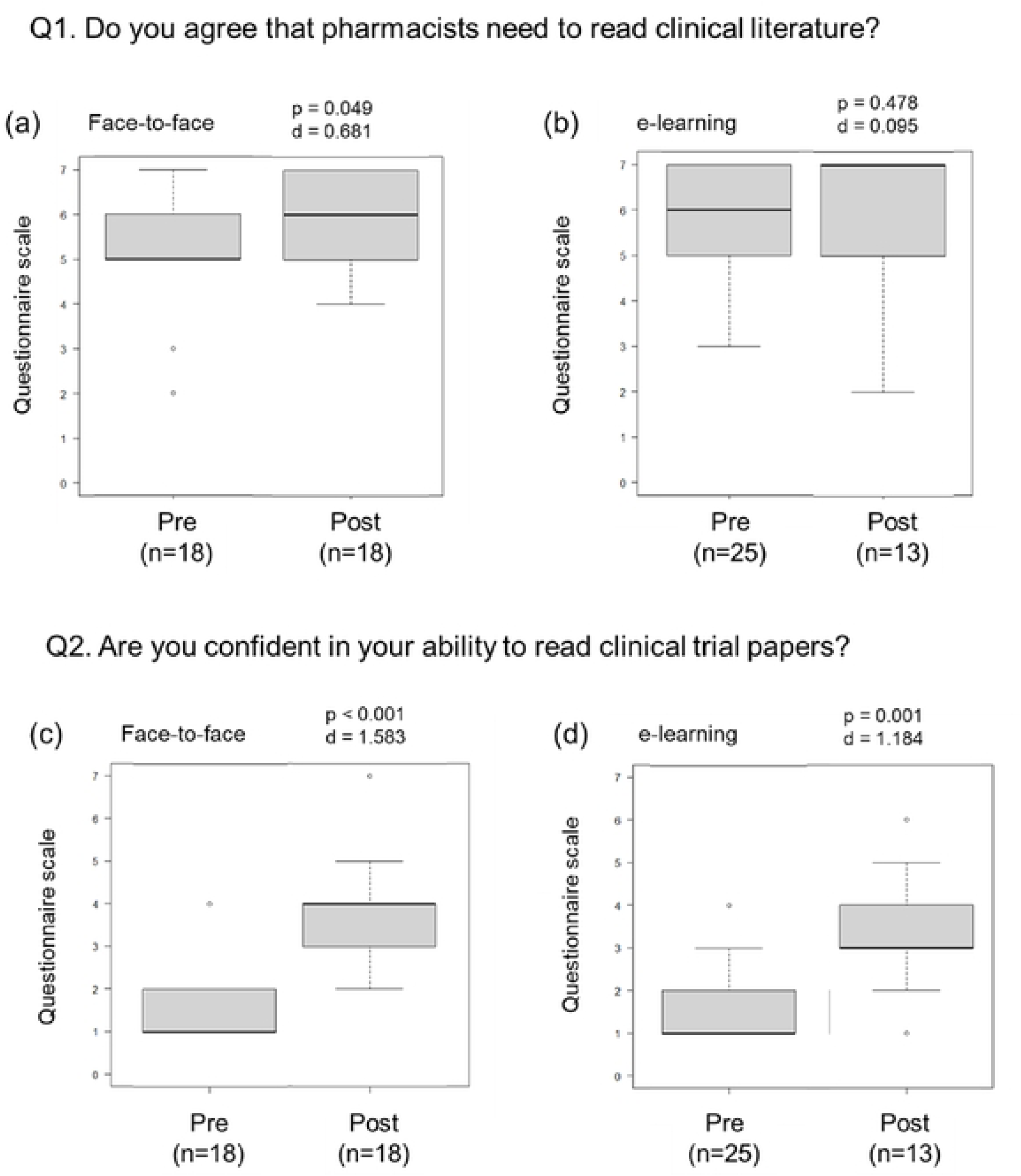
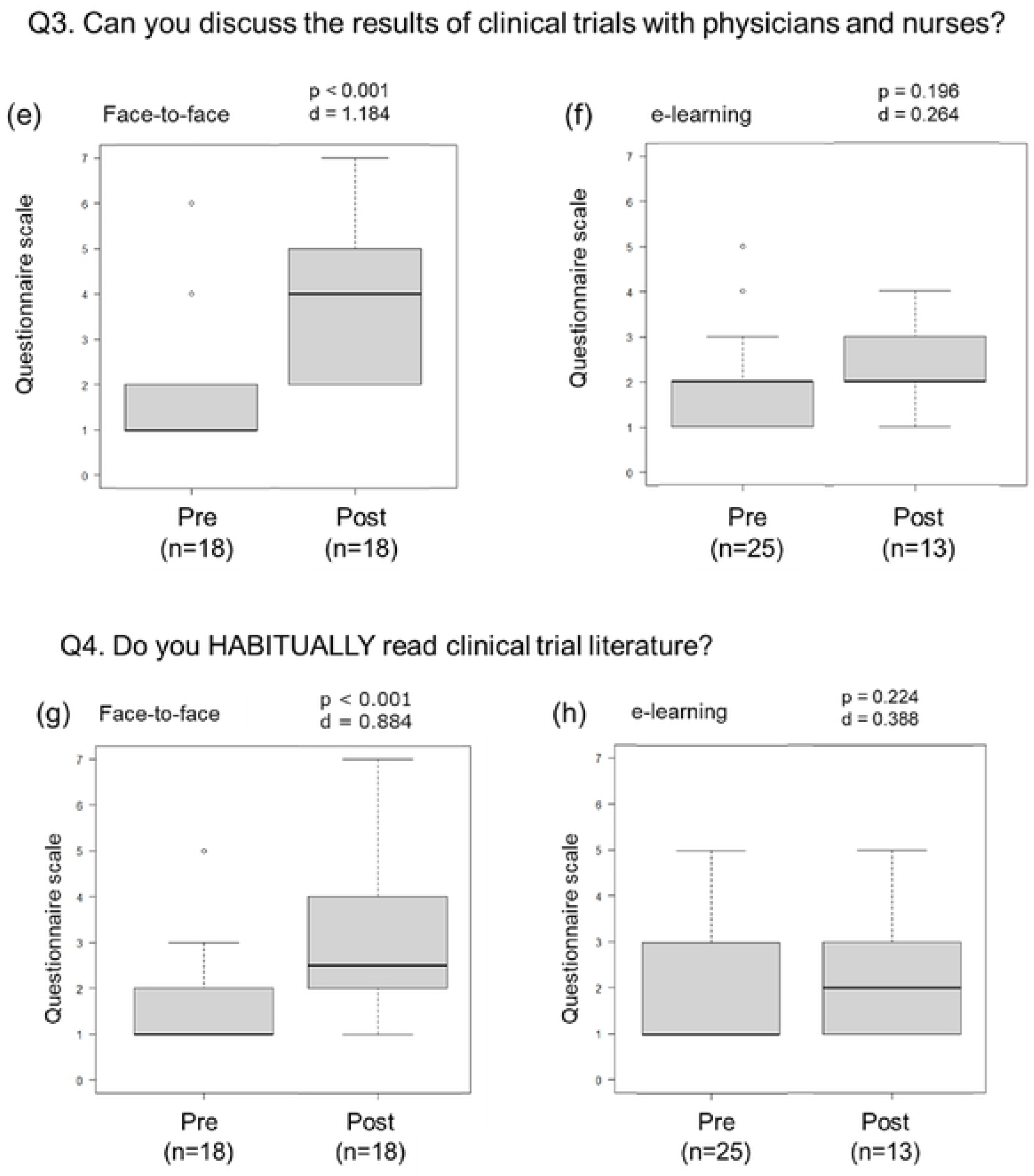
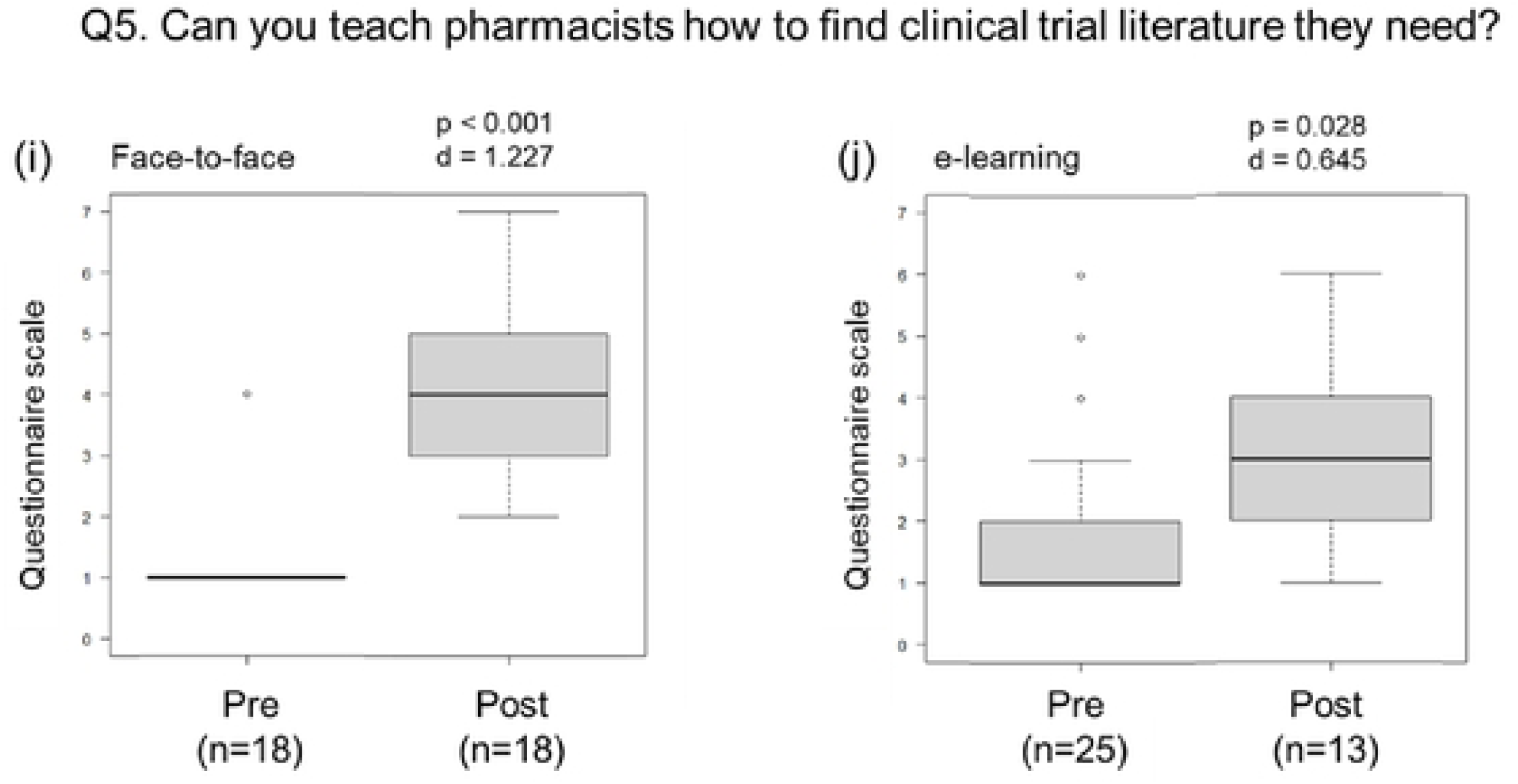
Results of questionnaire between face-to-face journal club and e-learning journal club. (a) QI. Do you agree that the pharmacists need to read clinical literature? (b) Q2. Are you confident in your ability to read clinical trial papers? (c) Q3. Can you discuss the results of clinical trials with physicians and nurses? (d) Q4. Do you HABITUALLY read clinical trial literature? (c) Q5. Can you teach pharmacists how to find clinical trial literature they need? (1: strongly disagree. 7: strongly agree)

## Discussion

The present study aimed to compare learning outcomes between the face-to-face journal club and the e-learning course. The study showed that the face-to-face journal club was able to produce significantly better learning outcomes than the e-learning course in terms of knowledge as well as awareness of critical appraisal of clinical literature.

Participants’ critical reading skills for clinical literature improved through attending the course in both groups. However, the face-to-face method was more effective than the e-learning method (Figure 1), suggesting that in-person interaction between instructors and participants can achieve more successful learning outcomes. Given that the preliminary knowledge of critical reading was significantly different between the two groups at baseline as indicated by the scores of pre-course exams, it would be necessary to compare the score growth to evaluate the outcomes. However, ceiling effects were observed in the face-to-face group, which precluded fair comparison. Nonetheless, the fact that a statistical difference between pre-course and post-course exam scores was observed in the face-to-face group, but not in the e-learning group, implies that learning outcomes in the face-to-face group were reliable.

Participants in both courses recognized the necessity of incorporating reading clinical literature into their regular duties as indicated by the relatively high scores (5 or 6) of the pre-course survey (Figure 2(a-b)). However, the score in the face-to-face group was even higher in the post-course survey, whereas there was no significant increase in the score in the e-learning group, which suggested that the face-to-face method can be more effective in promoting learners’ awareness of the need for critically reading clinical literature.

Less successful learning outcomes of the e-learning method might be attributed to its instructor-centered nature with little interaction between instructors and participants. The video-recorded e-learning modules were carefully constructed but the course could have been more effective by adopting a wider variety of interactive methods. For example, after mastering the basics of critical reading, the skills could be reinforced through periodic online discussion sessions.

Nonetheless, e-learning has some advantages. During the COVID-19 pandemic, remote learning methods including e-learning have often been utilized. In fact, our e-learning course was launched during the pandemic in order not to stop pharmacist education. It has been reported that dental students were satisfied with aspects of e-learning such as technical skills, accessibility to instructors and classmates, and improvement of oral communication skills [24]. Some researchers reported that e-learning seemed to be more effective than face-to-face training for the acquisition of theoretical knowledge for general medicine residents [25]. Additionally, an artificial intelligence (AI) integrated communication learning tool was created during the pandemic to educate pharmacy students [26]. When the need for social distancing returns to society, the results of these studies, including ours, have the potential to serve as a useful finding for educational activities in various fields. Effectiveness of remote learning methods including e-learning can depend on various factors. Therefore, this study does not reduce the potential of e-learning.

The present study did have limitations. First, significant difference in learning outcomes between the two methods may result from some confounding factors such as learning materials or contents of lectures. Further research is required where conditions other than learning methods are unified. Second, the dropout rate of the e-learning course was high (59%), which suggesting that participants may have found it difficult to implement the e-learning materials independently or that the e-learning materials might be too difficult for the participants. These results of our research have made it clear that when using e-learning materials in future, it is important to emphasize interactivity as much as possible to avoid the education becoming overly one-directional. The fact that the articles were written in English would also be associated with the high dropout rate. As machine translation tools have rapidly been more common and reliable, non-native speakers of English could make use of them for a quicker and better understanding of clinical literature.

## Conclusion

Learning outcomes of critical appraisal skills for the face-to-face journal clubs were significantly better than those for the online e-learning course. Although e-learning has advantage of not being subject to time or geographical limitations compared to face-to-face educational methods, its learning outcomes may be limited compared to face-to-face learning environments if the e-learning materials are difficult or if interactivity with participants cannot be established.

## Data Availability

All relevant data are within the manuscript and its Supporting Information files.

## Acknowledgments

We would like to express my deep appreciation to all the pharmacy students and pharmacists who attended the our face-to-face or online e-learning journal club.

## References

1. American College of Clinical Pharmacy. The definition of clinical pharmacy. Pharmacotherapy. 2008;28(6):816–817. PubMed PMID: 18503408.

2. Arif SA, Gim S, Nogid A, Shah B. Journal clubs during advanced pharmacy practice experiences to teach literature-evaluation skills. Am J Pharm Educ. 2012;76(5):88. PubMed PMID: 22761529.

3. Nakagawa N, Murai Y, Obara T, Ohara H, Kurita S, Lai L. Comparative Study of Pharmacists’ Perceptions of Clinical Literature Accessibility between Japan and USA –A Questionnaire Survey between Miyagi Prefecture and State of Florida–. Jpn J Drug Inform. 2018;19(4):180–187. DOI: 10.11256/jjdi.19.180.

4. Nakagawa N, Ishii Y, Sato Y, Suzuki H, Suzuki H, Murai Y. Verification of Outcomes of Journal Club as a Method of Improving Medical Terminology and Critical Reading Skills. Jpn J Pharm Health Care Sci. 2017;43(9):525–531. DOI: 10.5649/jjphcs.43.525.

5. Shimizu T, Ueda M, Toyoyama M, Ohmori S, Takagaki N. Evaluation of an Evidence-based Medicine Educational Program for Pharmacists and Pharmacy Students. Yakugaku Zasshi. 2017;137(8): 987–998. PubMed PMID: 28768952.

6. Ishii Y, Nakagawa N, Obara T, Ohara H, Kurita S, Murai Y. Survey on the Pharmacists Perceptions of Clinical Trial Literature Accessibility. Yakugaku Zasshi. 2020;140(9):1195–1198. PubMed PMID: 32879251.

7. Nakagawa N, Murai Y, Yoshida M, Suzuki H, Mano N. Effects of an evidence-based medicine workshop on Japanese pharmacy students’ awareness regarding the importance of reading current clinical literature. J Pharm Health Care Sci. 2015;1:23. PubMed PMID: 26819734.

8. Itani R, Khojah HMJ, Jaffal F, Rahme D, Karout L, Karout S. Provision of pharmaceutical care to suspected high-risk COVID-19 patients through telehealth: a nationwide simulated patient study. BMC Health Serv Res. 2021; 21(1):997. PubMed PMID: 34548092.

9. Sumiyoshi T, Morio Y, Kawashima T, Tachimori H, Hongo S, Kishimoto T et al. Feasibility of remote interviews in assessing disease severity in patients with major depressive disorder: A pilot study. Neuropsychopharmacol Rep. 2024;44(1):149–157. PubMed PMID: 38267023.

10. Norikoshi Y, Matsunaga Y, Uchida Y, Horio F, Anraku M, Suruki K et al. Changes in Attitudes to Medical Care among Pharmacy Pharmacists before and after the COVID-19 Pandemic: A Questionnaire Survey. Yakugaku Zasshi. 2023;143(12):1027–1038. PubMed PMID: 38044108.

11. Dyke EV, Jauncey-Cooke J, Johnston ANB. e-Learning interventions for nurses to prevent venous thromboembolism in patients: A realist review. J Clin Nurs. 2023;32(15-16):4441–4453. PMID: 36324243.

12. Papi A, Vestbo J, Fabbri L, Corradi M, Prunier H, Cohuet G et al. Extrafine inhaled triple therapy versus dual bronchodilator therapy in chronic obstructive pulmonary disease (TRIBUTE): a double-blind, parallel group, randomised controlled trial. Lancet. 2018;391(10125):1076–1084. PubMed PMID: 29429593.

13. Sher LD, Yiu G, Sakov A, Liu S, Caracta CF. Fluticasone propionate and fluticasone propionate/salmeterol multidose dry powder inhalers compared with placebo for persistent asthma. Allergy Asthma Proc. 2017;38(5):343–353. PubMed PMID: 28639542.

14. Wedzicha JA, Banerji D, Chapman KR, Vestbo J, Roche N, Ayers RT et al. Indacaterol-Glycopyrronium versus Salmeterol-Fluticasone for COPD. N Engl J Med. 2016;374(23):2222–2234. PubMed PMID: 27181606.

15. Byren I, Rege S, Campanaro E, Yankelev S, Anastasiou D, Kuropatkin G et al. Randomized controlled trial of the safety and efficacy of Daptomycin versus standard-of-care therapy for management of patients with osteomyelitis associated with prosthetic devices undergoing two-stage revision arthroplasty. Antimicrob Agents Chemother. 2012;56(11):5626–5632. PubMed PMID: 22908174.

16. Seemungal TA, Wilkinson TM, Hurst JR, Perera WR, Sapsford RJ, Wedzicha JA. Long-term erythromycin therapy is associated with decreased chronic obstructive pulmonary disease exacerbations. Am J Respir Crit Care Med. 2008;178(11):1139– 1147. PubMed PMID: 18723437.

17. Sethi S, Jones PW, Theron MS, Miravitlles M, Rubinstein E, Wedzicha JA et al. Pulsed moxifloxacin for the prevention of exacerbations of chronic obstructive pulmonary disease: a randomized controlled trial. Respir Res. 2010;11(1):10. PubMed PMID: 20109213.

18. Beasley R, Holliday M, Reddel HK, Braithwaite I, Ebmeier S, Hancox RJ et al. Controlled Trial of Budesonide-Formoterol as Needed for Mild Asthma. N Engl J Med. 2019;380(21):2020–2030. PubMed PMID: 31112386.

19. Nakagawa K, Garon EB, Seto T, Nishio M, Ponce Aix S, Paz-Ares L et al. Ramucirumab plus erlotinib in patients with untreated, EGFR-mutated, advanced non-small-cell lung cancer (RELAY): a randomised, double-blind, placebo-controlled, phase 3 trial. Lancet Oncol. 2019;20(12):1655–1669. PubMed PMID: 31591063.

20. Li HK, Rombach I, Zambellas R, Walker AS, McNally MA, Atkins BL et al. Oral versus Intravenous Antibiotics for Bone and Joint Infection. N Engl J Med. 2019;380(5):425–436. PubMed PMID: 30699315.

21. Kanda Y. Investigation of the freely available easy-to-use software ‘EZR’ for medical statistics. Bone Marrow Transplant. 2013;48(3):452–458. PubMed PMID: 23208313.

22. Faul F, Erdfelder E, Lang AG, Buchner A. G*Power 3: a flexible statistical power analysis program for the social, behavioral, and biomedical sciences. Behav Res Methods. 2007;39(2):175–191. PubMed PMID: 17695343.

23. Cohen J. A power primer. Psychol Bull. 1992;112(1):155–159. PubMed PMID: 19565683.

24. Syed J, Khan E, Kayal RA, Al Amoudi A, Nasir M, Hassan NN et al. Face-to-face and e-learning during the COVID-19 pandemic: A Pakistani dental undergraduates perspective. Work. 2024;78(3):551–558. PubMed PMID: 38277321.

25. Gaudin M, Tanguy G, Plagne M, Saussac A, Hansmann Y, Jaulhac B et al. E-learning versus face-to-face training: Comparison of two learning methods for Lyme borreliosis. Infect Dis Now. 2022;52(1):18–22. PubMed PMID: 34768016.

26. Nakagawa N, Odanaka K, Ohara H, Kisara S. Communication training for pharmacy students with standard patients using artificial intelligence. Curr Pharm Teach Learn. 2022;14(7):854–862. PubMed PMID: 35914846.

